# Leveraging large multi-center cohorts of Alzheimer Disease endophenotypes to understand the role of Klotho heterozygosity on disease risk

**DOI:** 10.1101/2021.10.07.21264646

**Authors:** Muhammad Ali, Yun Ju Sung, Fengxian Wang, Maria V. Fernández, John C. Morris, Anne M. Fagan, Kaj Blennow, Henrik Zetterberg, Amanda Heslegrave, Per M Johansson, Johan Svensson, Bengt Nellgård, Alberto Lleo, Daniel Alcolea, Jordi Clarimon, Lorena Rami, José Luis Molinuevo, Marc Suárez-Calvet, Estrella Morenas-Rodríguez, Gernot Kleinberger, Christian Haass, Michael Ewers, the Alzheimer’s Disease Neuroimaging Initiative (ADNI), Carlos Cruchaga

## Abstract

Two genetic variants (rs9536314 and rs9527025) in the Klotho gene, encoding a transmembrane protein, implicated on longevity and associated with brain resilience during normal aging, were recently shown to be associated with Alzheimer disease (AD) risk in cognitively healthy individuals that are APOE4 carriers. Specifically, the individuals heterozygous for this variant (KL-SV^HET+^), showed lower risk of developing AD. Furthermore, a neuroprotective effect of KL-VS^HET+^ has been suggested against amyloid burden for cognitively normal individuals. However, inconsistent associations and a smaller sample size of existing studies pose significant hurdles in drawing definitive conclusions. Here, we have performed a well-powered association analysis between KL-VS^HET+^ and five different AD endophenotypes; brain amyloidosis measured by positron emission tomography (PET) scans (n = 5,541) or cerebrospinal fluid Aβ levels (CSF; n = 5,093), as well as biomarkers associated with tau pathology: the CSF Tau (n = 5,127), phosphorylated Tau (pTau; n = 4,778) and inflammation: CSF soluble triggering receptor expressed on myeloid cells 2 (sTREM2; n = 2,123) levels. Our results found nominally significant associations of KL-VS^HET+^ status with biomarkers for brain amyloidosis (e.g. CSF Aβ positivity; odds ratio [OR] = 0.67 [95% CI, 0.55-0.78], β = 0.72, *P* = 0.007) and tau pathology (e.g. biomarker negative for CSF Tau; OR = 0.39 [95% CI, 0.19-0.77], β = -0.94, *P* = 0.007, and pTau; OR = 0.50 [95% CI, 0.27-0.96], β = -0.68, *P* = 0.04) in elderly (60-80 years old) individuals that are cognitively normal and APOE4 carriers. Our work supports previous findings and suggests that the KL-VS^HET+^ on a APOE4 genotype background may exert a protective effect by modulating the Aβ, Tau, and pTau burden and resulting cognitive decline in older controls susceptible to AD. The biological mechanism underlying APOE4 and KL-VS^HET+^ interaction and the neuroprotective effect of KL-related pathways against amyloid accumulation may warrant future investigation as a target for preclinical pharmacological studies to explore novel AD drug targets.

## Introduction

Alzheimer disease (AD), the most common form of dementia, affects about 30% of those aged over 85 years [1]. AD is classified as a neurodegenerative disease, affecting the brain integrity and functioning, eventually resulting in a progressive deterioration of cognitive capabilities [2]. Besides aging, a strong genetic risk factor for developing AD is the existence of epsilon 4 allele of the apolipoprotein E (APOE4) [3,4]. As such, subjects carrying APOE4 allele are significantly overrepresented among persons diagnosed with AD, in comparison to the non-carriers [5,6]. This particular genetic variant has been shown to be associated with cognitive decline [7] and reduced mean age at onset even within families with late onset AD [8]. Even among pre-symptomatic or cognitively normal subjects, carrying two copies of APOE4 allele (APOE4 homozygosity) has been reported to elevate the Aβ deposition [9–12], an earlier age-related memory decline [13], and an increased incidence of conversion [9], in comparison to APOE4 heterozygotes and noncarriers. These evidences suggest that APOE4 genotype lies at the core of AD pathophysiology, primarily due to its key role in cerebral Aβ pathology. Therefore, the search for potential genetic factors that interact with APOE4 genotype to reduce Aβ burden and eventually an individual’s risk for developing AD is crucial for halting the progression of disease and providing novel drug targets for an effective therapeutic intervention.

One such genetic factor that has been recently evaluated for its protective effect from developing AD in individuals that are cognitively normal and APOE4 carriers is Klotho (KL) protein heterozygosity [6,14,15]. KL is a transmembrane protein, implicated as a longevity factor [16,17] that promotes neuronal functions and brain resilience during aging [18–20]. In humans, two variants in the KL gene (rs9536314 for p.F352V and rs9527025 for p.C370S), that exist in a strong linkage disequilibrium, segregate together to form a functional haplotype called KL-VS. Interestingly, heterozygosity for the KL-VS haplotype (KL-VS^HET+^) has been associated with higher level of KL abundance in the serum [18,21], which in turn has been reported to exhibit protective effects such as healthy brain aging and protective synaptic functions, in comparison to the individuals that are carrying two copies of KL-VS haplotype (KL-VS^HET-^) [20,22]. Even though KL-VS^HET+^ is associated with better cognitive health and longevity among those aging normally, there exist no clear indication of its involvement in protection against aging-associated neurodegenerative disorders, such as AD.

Identification of genetic risk factors for AD based on clinical diagnosis includes several challenges. As such, AD diagnosis relies on the evidence of cognitive decline using standard cognitive tests that might be influenced by factors unrelated to disease e.g. anxiety, education, and general test-taking ability of the subject [23]. A complementary approach to the classical case-control studies is using intermediate phenotypes (endophenotypes) such as cerebrospinal fluid (CSF) biomarkers and Aβ burden assessed by positron emission tomography (PET) scanning. Using brain endophenotypes that are objective and highly reproducible, has the ability to provide enough statistical power to identify AD genetic risk factors, novel associations, and understand their impact on the brain [23,24]. For example, by using brain endophenotypes, researchers have identified novel protective genetic variants in *TMEM106B* and *MS4A* genes, associated with high neuronal proportion in AD [25], and increased CSF soluble triggering receptor expressed on myeloid cells 2 (*TREM2*) concentrations with reduced AD risk, respectively [26].

In the spectrum of AD pathology and different genetic factors that exert a protective effect in the context of disease onset and/or progression, KL-VS appears to be a compelling candidate due to its implication in promoting longevity and cognitive resilience during aging [18,20,22]. Interestingly, two recent studies aimed at accessing the protective effect of KL-VS^HET+^ against AD in cognitively normal individuals [15,27], and provided contradictory evidence. The first study [15] focused on 309 late-middle-aged adults (mean age 61 years) and found KL-VS^HET+^ to be associated with reduced Aβ aggregation, suggesting its protective effect against APOE4-linked pathways to disease onset in AD. The second study [27] focused on 581 adults (mean age 71 years) and found no significant associations between KL-VS^HET+^ and cognitive decline, independent of the APOE4 genotype, suggesting no modifying effect of KL-VS^HET+^ on Aβ aggregation and APOE4-driven cognitive decline in preclinical AD. Furthermore, a recent large-scale meta-analysis [6], focused on cognitively normal individuals in the age range of 60-80 years and revealed a 30% reduction in AD risk for subjects that are APOE4 carriers and KL-VS^HET+^. They also observed a significant association between KL-VS^HET+^ and higher Aβ42 in CSF (P-value 0.03) and lower Aβ on PET scans (P-value 0.04), modulated by APOE4 status. Due to contradictory outcomes from the existing reports and their relatively small sample sizes, we aimed at performing a systematic evaluation of association between KL-VS^HET+^ and multiple well-established AD endophenotypes to check whether it has a protective effect on AD. Here, we have performed a well-powered association analysis between KL-VS^HET+^ and five different AD endophenotypes; Aβ assessed by PET scans (n = 5,541) and CSF (n = 5,093), as well as the CSF Tau (n = 5,127), phosphorylated Tau (pTau; n = 4,778) and TREM2 (n = 2,123). In line with previous studies, we performed APOE4- and age-stratified (60-80 years) analyses, to determine if there is any association between KL-VS^HET+^ and Aβ aggregation that is modulated by APOE4 status. In addition, we also evaluated if there is any association between KL-VS^HET+^ and other AD endophenotypes that include Tau, pTau, and TREM2 measured by CSF. Briefly, in the case of APEO4-carriers, we found significant associations between KL-VS^HET+^ and biomarkers for brain amyloidosis (CSF Aβ42; *P*-value=0.007) and tau pathology (CSF Tau; *P*-value=0.007, and pTau; *P*-value=0.04). As evident from the observed *P*-values, the detected associations are nominally significant that would likely fail the multiple test correction, indicating the need of validating these findings in studies with even larger sample size for drawing definitive conclusion. Albeit nominally significant, these findings suggest that the combination of KL-VS^HET+^ and APOE4 genotype exert a protective effect by modulating the CSF Aβ, Tau, and pTau burden and resulting cognitive decline in older controls susceptible to AD.

## Methods

### Study samples and phenotype processing

For this study, we collected data from 17 different AD-related cohorts with a total sample size of 9,526 (Table S1). We analyzed the association between KL-VS^HET+^ and five different AD endophenotypes (Table 1) that served as biomarkers for brain amyloidosis (Aβ levels assessed by amyloid-PET [n = 5,541] and Aβ42 measured from CSF [n = 5,093]), tau pathology (Tau [n = 5,127] and pTau [n = 4,778] from CSF), and inflammation-specific (sTREM2 levels from CSF [n = 2,123]). These subjects come from Memory and Aging Project (MAP), Alzheimer’s Disease Neuroimaging Initiative (ADNI), BIOCARD, the Dominantly Inherited Alzheimer Network (DIAN), HB, Lleo, London, MOLI, Pau, Mayo Clinic (Mayo), SWEDEN, UPENN, UW, Progression Markers Initiative (PPMI), Anti-Amyloid Treatment in Asymptomatic Alzheimer’s Disease (A4), and ADNI Department of Defense (ADNIDOD) studies. A part of data used in the preparation of this article was obtained from the ADNI database (adni.loni.usc.edu). ADNI was launched in 2003 as a public-private partnership, led by Principal Investigator Michael W. Weiner. Collection of genotype data, PET image processing, and CSF data processing for each cohort are described in detail in the respective studies [10,11,25,26,28,29].

**Table 1.** Demographics of analyzed Alzheimer’s disease (AD) endophenotypes.

|  | **Total** | **Amyloid-**<br>**PET** | **Aβ42** | **Tau** | **pTau** | **TREM2** |
| --- | --- | --- | --- | --- | --- | --- |
| **Sample size** | 9,526 | 5,541 | 5,093 | 5,127 | 4,778 | 2,123 |
| **Female (%)** | 51.86 | 54.54 | 49.30 | 49.19 | 48.68 | 50.31 |
| **Age, mean (SD)** | 68.93 (11.13) | 69.53 (10.73) | 67.04 (13.27) | 67.15 (13.30) | 66.93 (13.43) | 68.17 (12.13) |
| **APOE4+ (%)** | 39.03 | 37.39 | 40.74 | 41.08 | 39.47 | 42.11 |
| **Biomarker, mean (SD)** | 0.028 (0.02) | 0.045 (1.03) | -0.0018 (1) | 0.027 (1) | 0.02 (1) | 0.05 (0.98) |
| **Klotho-VS<sup>HET+</sup> (%)** | 25.76 | 25.99 | 25.31 | 25.16 | 25.09 | 25.20 |
| **Cases** | 3,109 | 1,090 | 2,424 | 2,443 | 2,297 | 1,074 |
| **Controls** | 5,286 | 4,117 | 1,584 | 1,589 | 1,582 | 879 |

Demographics of subjects at the time of amyloid PET imaging and CSF sampling. This table summarizes basic demographic information of subjects included in the analysis. For each modality, we report percentage of females, mean age of the subjects and standard deviation (SD) in the age, percentage of *APOE4+* subjects, mean value of the endophenotypic biomarker and its SD, percentage of KL-VS heterozygous (KL-VS<sup>HET+</sup>) subjects, and number of cases and controls. To normalize endophenotypes across different cohorts, we converted different amyloid imaging measures (e.g. Centiloid, PiB, and AV45) into log-normalized z-score using “scale” function in base R. Phenotype from each cohort was normalized individually to account for within cohort variation. These AD endophenotypes are used for checking their association with KL-VS<sup>HET+</sup>. Abbreviations: PET, positron emission tomography; Aβ, β-amyloid; pTau, phosphorylated tau; soluble triggering receptor expressed on myeloid cells 2, TREM2; sd, standard deviation; KL-VS, Klotho-VS; Het+, heterozygosity.

Briefly, subjects were included if they were diagnosed as cognitively normal or AD, based on the clinical assessments from the respective studies. Any subject that was missing information about the sex, age, KL-VS^HET+^, APOE4 genotype, and genetic principal components (PCs), was excluded from the study. Following this rationale, we considered 3,725 cognitive normal individuals assessed by amyloid-PET and 1,030 cognitive normal individuals measured by CSF (AB42, Tau, pTau) and 639 individuals with CSF-TREM2 levels.

For each cohort, amyloid PET images were normalized to their reference cerebellar regions in order to obtain standardize uptake value ratios (SUVR) in a composite of cortical brain areas. The z-scores were calculated for each endophenotype using the mean and standard deviation (SD) units across each cohort and applied to the entire endophenotype.

All endophenotypes were also dichotomized as biomarker positive (case) or negative (control) using cutoffs defined for each endophenotype [11]. Defining biomarker positivity and negativity requires the selection of a cut-point at which to dichotomize. In order to dichotomize the scalar quantitative endophenotypes, we employed a gaussian mixture model (GMM) that relies on hierarchical model-based agglomerative clustering to get votes for defining a cut-point for dichotomization. We used Mclust function from “mclust” R package (version 5.4.6) for dichotomizing all endophenotype.

### Genotyping, quality checks, imputation, and population structure

We applied stringent quality control (QC) steps to process the genotyping array and sequencing data. We used the threshold of 98% for removing single nucleotide polymorphisms (SNPs) and individuals with low call rate. Autosomal SNPs that were not in the Hardy-Weinberg equilibrium (P < 1×10^−6^) were also removed. Subject duplication and relatedness was estimated from identity-by-descent (IBD) analysis carried out in Plink version 1.9 [30]. In case of related subjects (Pihat ≥0.25), the sample from MAP or with a higher number of variants that passed the QC was prioritized. For phasing and imputation, we used The 1000 Genomes Project Phase 3 data (October 2014), SHAPEIT v2.r837 [31], and IMPUTE2 v2.3.2 [32]. We used impute probability score < 0.90 and ≥0.90 as thresholds for missing and fully observed individual genotypes, respectively. Genotyped and imputed variants with MAF < 0.02 or IMPUTE2 information score < 0.30 were discarded. Principle component analysis (PCA) was performed on the genotype data to obtain genetic PCs that capture population substructure (supplementary figure S2). In order to obtain the largest and most homogeneous pool of population, only European individuals were considered (supplementary figure S3) for the subsequent statistical analyses.

### Statistical Analyses

Statistical analyses and data visualization were performed in Plink version 1.9 [30] and R version 3.5.2 [33]. We performed association analyses of KL-VS^HET+^ status with different AD endophenotypes from PET scan (Aβ) and CSF (Aβ42, Tau, pTau, and TREM2). The associations between the biomarker positivity and KL-VS^HET+^ were tested using general linear model (GLM). The implementation of GLM from the base R [33] “stats” package was used for the evaluation of association and the outcomes measurements were adjusted for sex, age, and first three genetic PCs. For the Aβ levels measured by PET scan, we considered the age at scan and in the case of endophenotype levels measured by CSF, we considered subject age when lumbar puncture was performed. Furthermore, associations were evaluated across three different strata, (1) all subjects (AD and controls), (2) subjects aged 60 to 80 years (AD and controls), and (3) only control subjects aged 60 to 80 years. All association analyses were stratified by APOE4 status, APOE4 carriers (APOE-24, 34, 42, 43, and 44) and APOE4 non-carriers (APOE-22, 23, 32, and 33). Associations were deemed significant at a threshold of *P*-value < 0.05. A schematic overview of conducted analyses and used datasets is provided in Figure 1.

**Figure 1:**
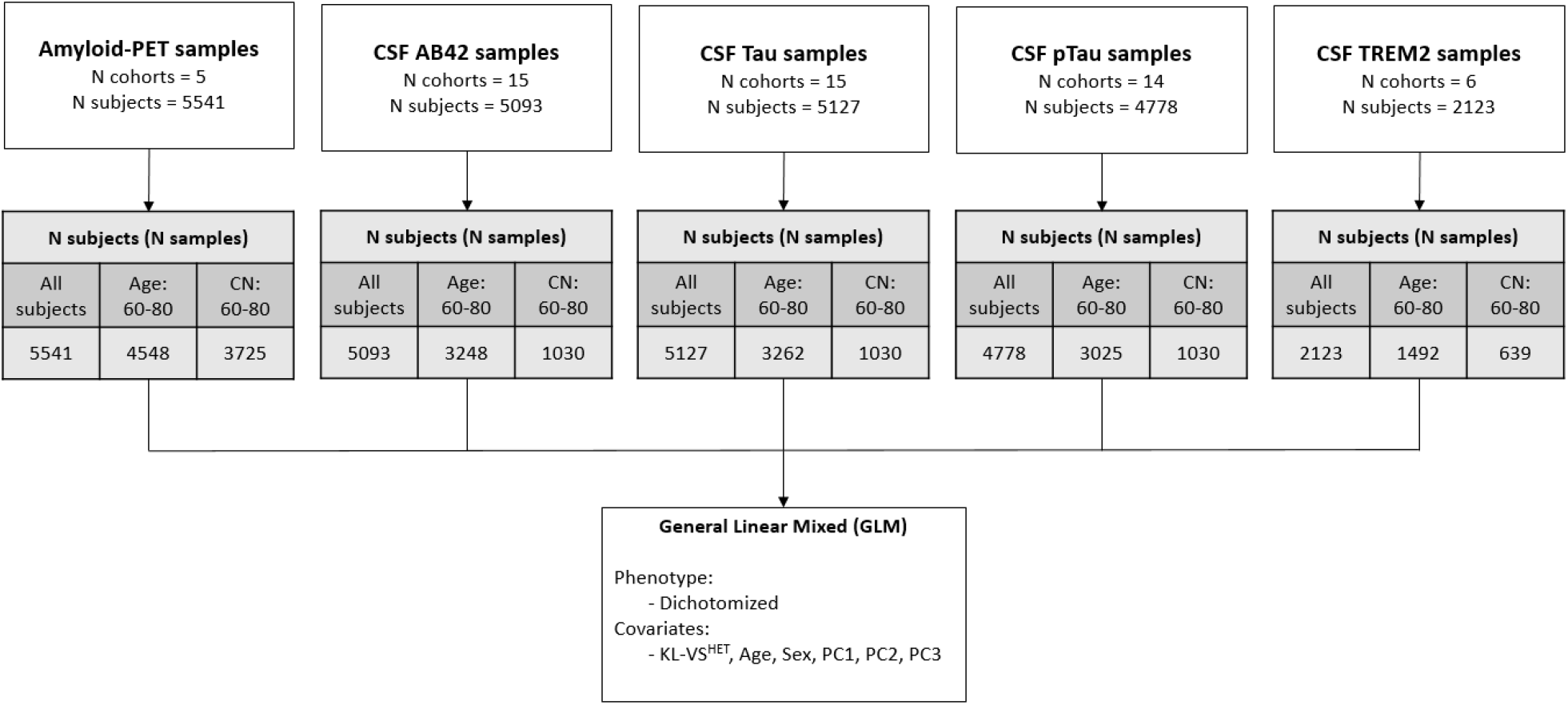
Schematic overview of datasets and performed analysis. Schematic overview of datasets and performed analysis. Number of subjects in each modality were stratified into three categories: 1) All of the subjects; 2) Age: 60-80, Subjects aged 60 to 80 years; 3) CN: 60-80, Control subjects aged 60 to 80 years. Association between KL-VS^HET^ and endophenotypes were assessed using multiple linear regression model for dichotomized phenotype. Age, Sex, and first three genetic PCs were used as covariates in an *APOE*4-stratified analysis. Abbreviations: PET, positron emission tomography; N, number of; CSF, cerebrospinal fluid; Aβ, β-amyloid; pTau, phosphorylated tau; CN, controls; KL, Klotho; Het, heterozygous; PC, principal component.

## Results

The aim of this study was to determine whether there is a significant association between KL-VS^HET+^ and AD endophenotypes in cognitively normal individuals that are APOE4-carriers. To accomplish this goal, we analyzed genetic data and five different AD endophenotypes from 9,526 individuals obtained through 17 different AD-related cohorts. In line with existing studies [6,15], our findings validate the protective effect of KL-VS^HET+^ against AD in healthy individuals that are APOE4 carriers.

### Association between KL-VS^HET+^ status and brain amyloidosis measured by PET scan and CSF

We evaluated the association of KL-VS^HET+^ status against Aβ levels measured by the PET scan and CSF for 5,541 and 5,093 subjects, respectively (Table 1). Although associations were evaluated for three different age ranges (Table S2), our main focus was control subjects that are 60 to 80 years old (Table 2). We focused on this age range for being consistent with existing studies that report a pronounced effect of APOE4 positivity in AD between age 60 to 80 years in comparison to older individuals (≥80 years) [6,34]. While KL-VS^HET+^ status was associated with decreased levels of Aβ positivity observed through the PET scan, there was no significant association found across any age group or APOE4 stratification (Table S2). On the other hand, we found a significant association between KL-VS^HET+^ status and CSF Aβ biomarker positivity in cognitively normal APOE4+ individuals that are 60 to 80 years old (Table 2; OR = 0.67 [95% CI, 0.55-0.78], β = 0.72, *P* = 0.007). We also observed a significant association for APOE4 non-carriers, however, the effect size and the strength of the association was lower in this group (OR = 0.61 [95% CI, 0.51-0.70], β = 0.46, *P* = 0.03) than the APOE4 carriers (Table 2). Taken together, we were able to replicate the previously reported associative findings [6,34] between increased Aβ CSF levels and KL-VS^HET+^ in a larger sample group.

**Table 2.** Genetic association of KL-VSHET+ with AD endophenotypes in cognitively normal subjects aged 60–80 years, stratified by APOE4 status.

| Modality | Group | CN subjects<br>(KL-VS<sup>HET+</sup> %) | Odds ratio | Estimate | P value |
| --- | --- | --- | --- | --- | --- |
| **Amyloid PET** | APOE4+ | 1328 (27.5) | 0.94 | -0.07 | 0.61 |
|  | APOE4- | 2397 (26.5) | 0.99 | -0.01 | 0.90 |
| **Aβ42** | APOE4+ | 308 (26.3) | 0.67 | 0.72 | **0.007** |
|  | APOE4- | 722 (26.3) | 0.61 | 0.46 | **0.03** |
| **Tau** | APOE4+ | 308 (26.9) | 0.39 | -0.94 | **0.007** |
|  | APOE4- | 722 (26.5) | 0.85 | -0.16 | 0.49 |
| **pTau** | APOE4+ | 308 (26.9) | 0.50 | -0.68 | **0.04** |
|  | APOE4- | 722 (26.3) | 0.89 | -0.11 | 0.61 |
| **TREM2** | APOE4+ | 199 (31.2) | 1.08 | 0.08 | 0.80 |
|  | APOE4- | 440 (25.2) | 1.20 | 0.18 | 0.43 |

Association between KL-VS<sup>HET+</sup> and different dichotomized AD endophenotypes were assessed using general linear model (GLM). We used dichotomized endophenotype as the response variable, whereas, age, sex, and first three genetic PCs were used as covariates in an *APOE4*-stratified analysis. Significant associations are represented by bold P-values. Abbreviations: KL-VS<sup>HET+</sup>, Klotho-VS heterozygous; CN, Control; AD, Alzheimer's disease; Std. Error, Standard error; %, percentage; Aβ, β-amyloid; pTau, phosphorylated tau; soluble triggering receptor expressed on myeloid cells 2, TREM2.

### APOE4-related alteration in tau pathology varies by KL-VS^HET+^ status

Similar to Aβ association analyses, we also determined if there was an age- and APOE4-dependent association of KL-VS^HET+^ status with dichotomized CSF Tau and pTau levels. In the age range of 60 to 80 years, we observed significant association between KL-VS^HET+^ status and CSF Tau (OR = 0.39 [95% CI, 0.20-0.77], *P* = 0.007), and pTau (OR = 0.50 [95% CI, 0.27-0.96], *P* = 0.04) levels in elderly (60-80) cognitive normal APOE4+ participants (Table 2). In both cases, the KL-VS^HET+^ was associated with being biomarker negative (e.g., lower CSF Tau/pTau levels which are associated with lower AD risk (β = -0.94 and -0.68 for Tau and pTau, respectively). Regarding the effect size, we observed an almost 6-fold decrease for both Tau and pTau levels in APOE4 carriers as compared to the non-carriers, suggestive of a more pronounced protective effect for subjects carrying one copy of KL-VS haplotype. As expected, a consistent negative association was observed for both modalities when effect sizes were represented in the form of a Forest plot (Figure 2). While we observed a consistent effect of KL-VS^HET+^ on tau pathology biomarker negativity for controls subjects that are APOE4 non-carriers (β = -0.16 and -0.11 for Tau and pTau, respectively), the association was not deemed significant in this stratum (*P* = 0.49 and 0.61 for Tau and pTau, respectively).

**Figure2:**
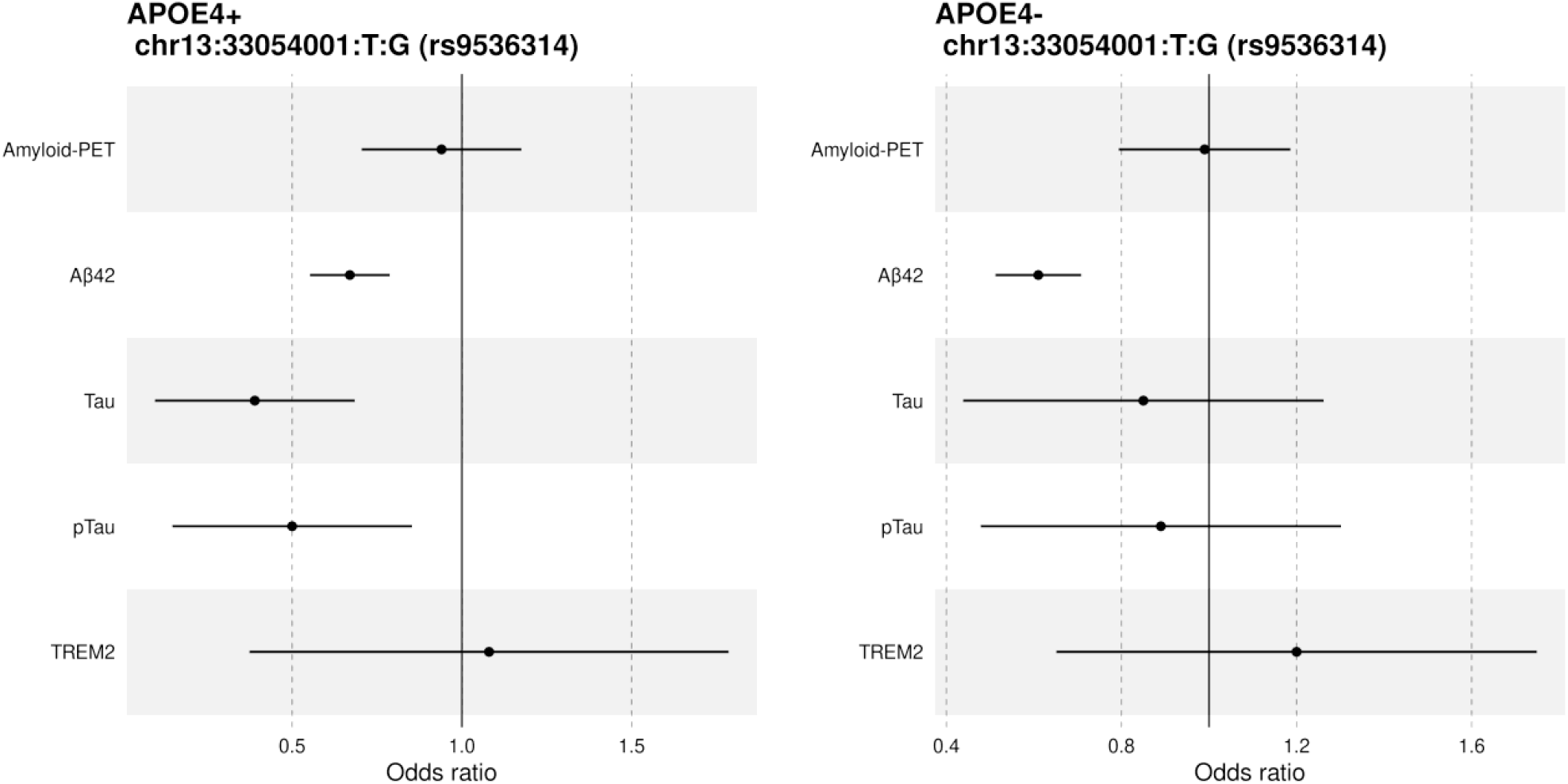
Forest plot of odds ratio (OR) for KL-VS^HET+^ association with dichotomized AD endophenotypes in 60-80 year control subjects, stratified by *APOE*4 status. A significant association was detected between KL-VS^HET+^ and dichotomized Aβ, Tau, and pTau CSF levels. In case of Aβ, the associations ware deemed significant across both APOE strata, whereas for Tau and pTau, associations were observed only in APOE4 carriers, representing an exclusive protective effect of KL-VS^HET+^ for the cognitively normal individuals who are aged 60 to 80 years and carrying APOE4. Abbreviations: *APOE*4+, *Apolipoprotein E*4 positive; *APOE*4-, *Apolipoprotein E*4 negative; PET, positron emission tomography; Aβ, β-amyloid; pTau, phosphorylated tau.

### Direct effect of KL-VSHET^+^ status on inflammation-specific biomarker

For TREM2 CSF levels, we did not observe any significant association between this inflammation-specific biomarker and KL-VS^HET+^ status across any subject stratification, regardless of the age group and APOE4 status. Nevertheless, for control subjects aged 60 to 80 years, we observed that KL-VS^HET+^ status is associated with increased TREM2 CSF levels, which represents the positivity of inflammation-specific biomarker, but association was not deemed significant in APOE4 carriers (OR = 1.08 [95% CI, 0.58-2], β = 0.08, *P* = 0.80) as well as non-carriers (OR = 1.20 [95% CI, 0.77-1.86], β = 0.18, *P* = 0.43). These findings suggest the protective effect of KL-VS^HET+^ status by modulating the CSF TREM2 levels as existing literature has also associated increased CSF TREM2 levels with lower AD risk and slower progression [26,29].

### Sensitivity analysis result: Robustness of association between KL-VS^HET+^ status and brain amyloidosis to the APOE4 status, age, and sex

In order to estimate whether association between KL-VS^HET+^ and amyloidosis are robust to uneven sample size of APOE4 carriers and non-carriers as well as differences in the sex and age of the participants, the association analysis was repeated with same number of APOE4 carriers (N = 308) and non-carriers that are age- and sex-matched. As in the full-sample analyses for control subjects aged 60 to 80, the age-, sex-, and APOE4-group matched sample analyses revealed that, for KL-VS^HET+^, APOE4 status was consistently associated with CSF Aβ biomarker positivity (OR = 0.68 [95% CI, 0.56-0.78], β = 0.75, *P* = 0.005; Table S3). In accordance with our findings from full-sample analysis, we also observed a significant association in case of APOE4 non-carriers, however, the effect size and the strength of the association was also lower in this group (OR = 0.69 [95% CI, 0.51-0.79], β = 0.70, *P* = 0.034) than the APOE4 carriers. Moreover, similar trends were observed in the case of amyloid imaging, CSF Tau, pTau, and TREM2 in age- and sex-matched as well as the full-sample analyses for control subjects aged 60 to 80 (Table S3). We have observed that correcting for class imbalance of APOE4-carriers and non-carriers as well as for males and females of same age, the direction of effect remains the same for each endophenotype, however, strength of association becomes more profound (lower p-values). Taken together, these results suggest that observed associations between KL heterozygous control individuals and different AD endophenotypes is independent of the distribution of APOE4 carriage status, age, and sex of participants.

### Cognitively normal subjects (aged 60 to 80) drive association between KL-VS^HET +^ status and AD endophenotypes

We also conducted an APOE4-stratified association analyses between KL-VS^HET+^ status and biomarkers for brain amyloidosis (Aβ from PET and CSF), tau pathology (CSF Tau and pTau), and inflammation (CSF TREM2) for all participants, regardless of their age and case-control status, and no significant association was observed in any of the five endophenotypes (Table S2). Similarly, we also performed the same analyses for elderly samples that are 60-80 years old, regardless of their case-control status, and even in that case no significant association was detected across any endophenotype (Table S2). Nevertheless, it was interesting to see how the effects became apparent when we stratified the participants to the age range 60-80 and considered only the cognitively normal subjects, as in that case a significant association was detected between KL-VS^HET+^ status and CSF Aβ, Tau, and pTau levels (Table S2). Notably, these findings suggest that the detected nominally significant associations were mainly driven by the control participants that are 60 to 80 years old.

## Discussion

The role of Klotho protein as a longevity factor is widely recognized [16,17]. There has been an increasing amount of evidence supporting the relationship between KL-VS^HET+^ and preserved brain integrity and cognitive performance during normal aging [18–20]. In this study, we examined the association of KL-VS^HET+^ status with five different AD-associated endophenotypes assessed from CSF biomarkers (Aβ, Tau, pTau, TREM2) and PET scans (Aβ). Specifically, we checked for association between KL-VS^HET+^ status and biomarkers for brain amyloidosis (Aβ levels measured from CSF and amyloid PET) and neurodegeneration (CSF Tau and pTau). To our knowledge, we have analyzed the largest sample size of AD endophenotypic data for evaluating its association with KL-VS^HET+^, that is instrumental to discern the potential protective effect of this heterozygous genetic variant for AD in control APOE4 carriers. Our results showed that KL-VS^HET+^ status was associated with CSF Aβ and Tau/pTau biomarker negativity in individuals that are cognitively normal APOE4 carriers within age range of 60 to 80 years, suggesting that KL-VS^HET+^ status reduces the risk of AD in APOE4 carriers by lowering the AD pathology burden [6,34].

We were able to replicate the findings by Belloy et al [6]; that is, KL-VS^HET+^ status was significantly associated with increased CSF Aβ levels (Aβ biomarker positivity) for control subjects aged 60 to 80 years that are APOE4 carriers (OR = 0.67 [95% CI, 0.55-0.78], β = 0.72, P = 0.007). This association was also significant in case of APOE4 non-carriers (OR = 0.61 [95% CI, 0.51-0.70], β = 0.46, *P* = 0.03), however, the effect and significance was lower suggesting a stronger protective interaction between APOE4 carriers and KL-VS^HET+^ status (Table 2). Although, no significant association is observed in the case of Aβ from PET, the detected negative association is suggesting the protective effect of KL-VS^HET+^ by reducing the Aβ deposition, contrary to its higher levels that are found to be closely related to Aβ deposition in neuritic plaques [35] - a hallmark of AD. Discordance in Aβ PET and CSF biomarkers has already been shown to have important consequences for their application in inspectional, clinical, and trial settings [36]. For example, a previous study investigating longitudinal differences in cognition between participants without dementia with different CSF and PET profiles found no memory decline in concordant-negative (CSF−/PET−) and discordant (CSF+/PET−) groups, in contrast to the concordant-positive (CSF+/PET+) group that deteriorated over time [37]. Furthermore, it is highly likely that observed discordance between Aβ levels from PET and CSF is because changes in CSF Aβ levels occur before detectable alterations in amyloid PET during preclinical stages of AD. Palmqvist et al [36] reported similar results, when they analyzed 437 non-demented subjects from ADNI that had Aβ levels from PET scans and CSF, showing that CSF Aβ levels becomes abnormal in the earliest stages of AD, before amyloid PET and before neurodegeneration starts

We also investigated whether KL-VS^HET+^ status is significantly associated with Tau and pTau burden assessed from CSF. We found that KL-VS^HET+^ status was significantly associated with decreased Tau (OR = 0.39 [95% CI, 0.20-0.77], β = -0.94, *P* = 0.007) and pTau (OR = 0.50 [95% CI, 0.27-0.96], β = -0.68, *P* = 0.04) levels (CSF Tau/pTau biomarker negativity) in subjects that are APOE4 carriers and 60 to 80 years old. Interestingly, in the case of APOE4 non-carriers, albeit having the similar negative trend, the associations were not significant for both Tau (OR = 0.85 [95% CI, 0.54-1.35], β = -0.16, *P* = 0.49) and pTau (OR = 0.89 [95% CI, 0.57-1.39], β = -0.11, *P* = 0.61). This indicates that KL-VS^HET+^ status interaction with pathological aspects of AD are more profound in APOE4 carriers, such as Aβ and Tau accumulation during the pre-clinical phase of the disease [38,39]. Indeed, previous studies have also reported a significant association between KL-VS heterozygosity and reduced tau accumulation and lower memory impairment in elderly humans at risk of AD dementia [40,41]. However, in a mouse model of AD that was used to examine the neuroprotective effects of Klotho protein against neuronal damage associated with oxidative stress and neurodegeneration, no changes in Tau phosphorylation were observed in the presence of Klotho [42]. Unlike Tau and pTau association with KL-VS^HET+^ status, we observed a positive association in the case of CSF levels of soluble TREM2 (inflammation-specific biomarker). However, the observed increase in the CSF TREM2 levels was not significantly associated with KL-VS^HET+^ status for both APOE carriers (OR = 1.08 [95% CI, 0.59-2], β = 0.08, *P* = 0.80) and non-carriers (OR = 1.20 [95% CI, 0.77-1.86], β = 0.18, *P* = 0.43). Interestingly, recent studies [26,29] have shown that higher TREM2 levels are associated with lower AD risk and slower progression. Therefore, the observed positive association suggests that the protective effect of the KL-VS^HET+^ might be mediated by higher CSF TREM2 levels. However, this hypothesis will need to be validated in studies with larger sample size for CSF TREM2 levels.

Taken together, the observed significant associations between KL-VS^HET+^ status and biomarkers for brain amyloidosis (CSF Aβ positivity) and neurodegeneration (CSF Tau and pTau negativity) are suggestive of neuroprotective effect of KL-VS^HET+^ against age-related biomarker, biomolecular, and cognitive alterations that confer risk for AD. Our results further strengthen the findings by a recent meta-analysis including 25 independent studies, showing that APOE4 carriers who were also KL-VS^HET+^, were at a reduced risk for the combined outcome of conversion to mild cognitive impairment (MCI) or AD [6].

Notably, we assessed the associations between KL-VS^HET+^ status and AD endophenotypes across three age strata; all of the subjects (AD and controls), only subjects aged 60 to 80 years (AD and controls), and only control subjects aged 60 to 80 years (Table S2). Owing to a higher genetic risk for AD attributable to APOE4 in individuals that are 60 to 80 years old [43–45] and an existing study that hypothesized protective association of KL-VS^HET+^ status to be strongest in APOE4 carriers who are 60 to 80 years old [6], the focus of the current study was also at this particular age range. Although we observed similar associative trends most of the time, it was interesting to see how the effects became apparent when stratifying the cognitively normal subjects to the age range of 60 to 80 years. In all of the cases, no significant associations were observed between KL-VS^HET+^ status and AD endophenotypes while considering all of the AD and control subjects, or all of the AD and control subjects within age range of 60-80. However, pronounced effects and associations were apparent for Aβ, Tau, and PTau levels from CSF, while considering control individuals that are APOE4 carriers and KL-VS^HET+^. These findings suggest that the control group aged 60 to 80 years mainly drove the outcome in our analyses, further strengthening the existing hypothesis that KL-VS heterozygous genotype is favorable for survival in older people as compared to the young adults [46]. Importantly, we also observed that associations between KL heterozygous control individuals and different AD endophenotypes are robust to uneven sample size of APOE4 carriers and non-carriers as well as differences in the sex and age of the participants (Table S3). Although, the observed findings are in line with our initial hypothesis and confirming existing literature [6,34], the detected associations are nominally significant that would likely fail the multiple test correction due to limited sample size. Therefore, additional studies are required to investigate the associations between KL-VS^HET+^ and AD endophenotypes with relatively larger sample size to draw definitive conclusions.

The exact mechanism behind KL-VS^HET+^ interaction with APOE4 and modulation of Aβ, Tau, and pTau burden is yet unknown. However, it is logical to presume that KL-VS^HET+^ may confer resilience by increasing the serum level of circulating Klotho protein [18,21] or by changing its function. In animal mouse models, elevated klotho levels have led to an extended lifespan [17], enhanced cognition [19] and increased resilience to AD-related toxicity [47]. Other studies in humans, indicated that KL-VS^HET+^ status has protective effects against brain aging and cognitive decline [21,46], suggestive of its protective association against AD. Our findings also suggest that middle-aged APOE4 carriers that are KL-VS^HET+^ might be biologically younger than those who are APOE4 non-carriers and KL-VS^HET-^, and thus show resilience to age-induced cognitive and tau changes. Interestingly, we have observed an age-specific association between KL-VS^HET+^ and AD endophenotypes, which is in line with existing studies reporting a specific time window for the effect of KL-VS polymorphism [20,46].

To conclude, our work contribute to the existing state of the literature by demonstrating that the protective effects of KL-VS^HET+^ extend to age-related Aβ, Tau, and pTau burden and deficits in memory and executive function in control APOE4 carriers susceptible for AD. One promising research avenue for the future studies could be to assess whether Klotho protein levels in the CSF or serum of individuals associate with measures of AD and preclinical disease.

## Data Availability

The data will be made public upon acceptance of this article in a peer-reviewed journal.

## Acknowledgements

ADNI acknowledgement: Data collection and sharing for this project was funded by the Alzheimer’s Disease Neuroimaging Initiative (ADNI) (National Institutes of Health Grant U01 AG024904) and DOD ADNI (Department of Defense award number W81XWH-12-2-0012). ADNI is funded by the National Institute on Aging, the National Institute of Biomedical Imaging and Bioengineering, and through generous contributions from the following: AbbVie, Alzheimer’s Association; Alzheimer’s Drug Discovery Foundation; Araclon Biotech; BioClinica, Inc.; Biogen; Bristol-Myers Squibb Company; CereSpir, Inc.; Cogstate; Eisai Inc.; Elan Pharmaceuticals, Inc.; Eli Lilly and Company; EuroImmun; F. Hoffmann-La Roche Ltd and its affiliated company Genentech, Inc.; Fujirebio; GE Healthcare; IXICO Ltd.; Janssen Alzheimer Immunotherapy Research & Development, LLC.; Johnson & Johnson Pharmaceutical Research & Development LLC.; Lumosity; Lundbeck; Merck & Co., Inc.; Meso Scale Diagnostics, LLC.; NeuroRx Research; Neurotrack Technologies; Novartis Pharmaceuticals Corporation; Pfizer Inc.; Piramal Imaging; Servier; Takeda Pharmaceutical Company; and Transition Therapeutics. The Canadian Institutes of Health Research is providing funds to support ADNI clinical sites in Canada. Private sector contributions are facilitated by the Foundation for the National Institutes of Health (www.fnih.org). The grantee organization is the Northern California Institute for Research and Education, and the study is coordinated by the Alzheimer’s Therapeutic Research Institute at the University of Southern California. ADNI data are disseminated by the Laboratory for Neuro Imaging at the University of Southern California. HZ is a Wallenberg Scholar supported by grants from the Swedish Research Council (#2018-02532), the European Research Council (#681712), Swedish State Support for Clinical Research (#ALFGBG-720931), the Alzheimer Drug Discovery Foundation (ADDF), USA (#201809-2016862), the AD Strategic Fund and the Alzheimer’s Association (#ADSF-21-831376-C, #ADSF-21-831381-C and #ADSF-21-831377-C), the Olav Thon Foundation, the Erling-Persson Family Foundation, Stiftelsen för Gamla Tjänarinnor, Hjärnfonden, Sweden (#FO2019-0228), the European Union’s Horizon 2020 research and innovation programme under the Marie Skłodowska-Curie grant agreement No 860197 (MIRIADE), and the UK Dementia Research Institute at UCL.

KB is supported by the Swedish Research Council (#2017-00915), the Alzheimer Drug Discovery Foundation (ADDF), USA (#RDAPB-201809-2016615), the Swedish Alzheimer Foundation (#AF-742881), Hjärnfonden, Sweden (#FO2017-0243), the Swedish state under the agreement between the Swedish government and the County Councils, the ALF-agreement (#ALFGBG-715986), the European Union Joint Program for Neurodegenerative Disorders (JPND2019-466-236), the National Institute of Health (NIH), USA, (grant #1R01AG068398-01), and the Alzheimer’s Association 2021 Zenith Award (ZEN-21-848495).

---

**Figure S1.**
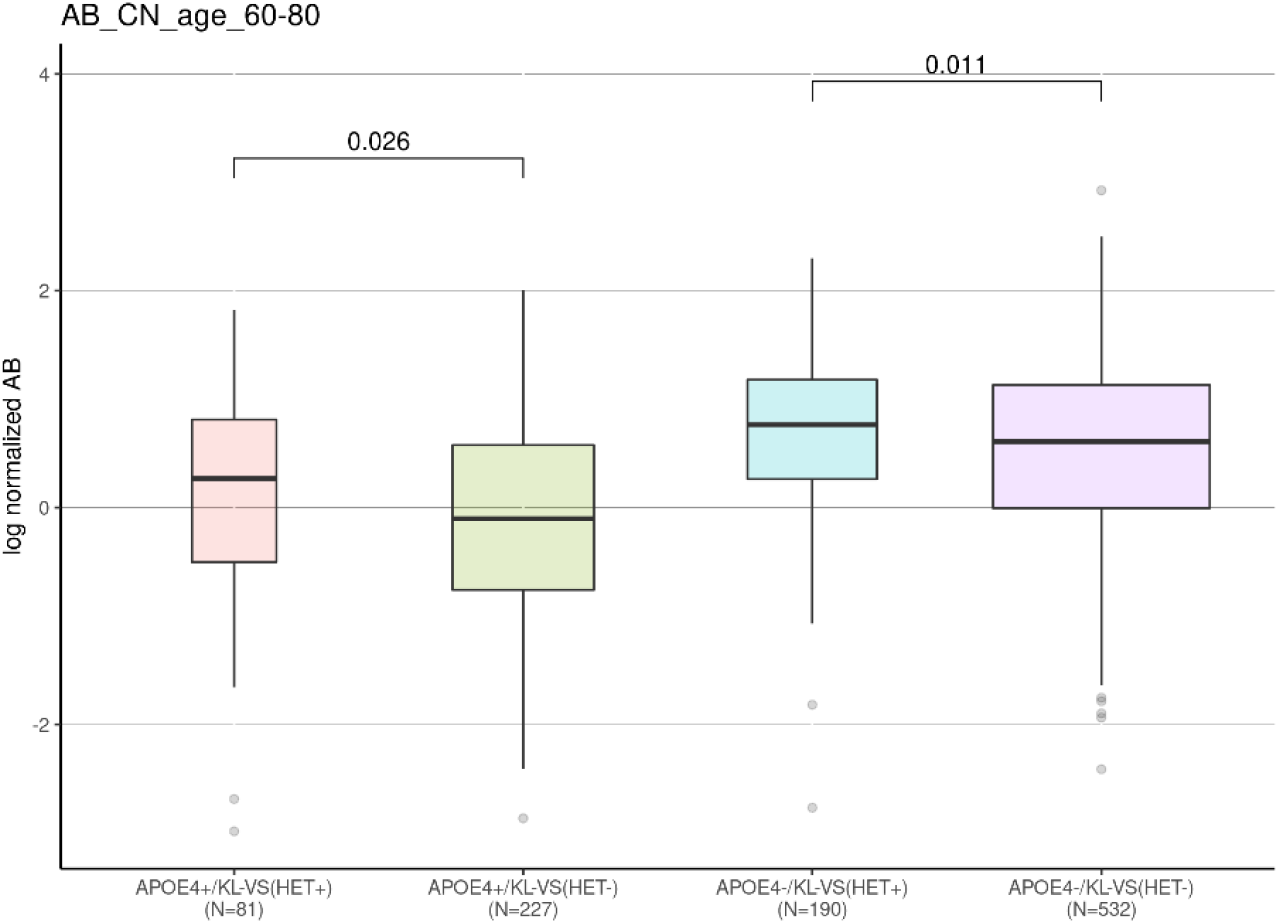
KL-VS heterozygosity status is associated with CSF amyloid beta levels in cognitively normal individuals. A significant association was detected between Klotho-VS^HET+^ and dichotomized CSF amyloid beta (Aβ) levels. The distribution of quantitative log-normalized CSF Aβ levels are shown in this box plot, stratified by *APOE*4 and KL-VS^HET+^ status, where width of the boxplot represents the sample size of each group. Box plot error bars show the 95th-percentile range. Gray circles indicate values outside of the 95th percentile range. There is a significant difference between the means of compared groups (*APOE*4+/ KL-VS^HET+^ vs. *APOE*4+/ KL-VS^HET-^ and *APOE*4-/ KL-VS^HET+^ vs. *APOE*4-/ KL-VS^HET-^) with mean comparison p-values labelled on the top. Abbreviations: *APOE*4+, *Apolipoprotein E*4 positive; KL-VS^HET+^, Klotho-VS heterozygous; KL-VS^HET-^, Klotho-VS homozygous; N, number of samples.

**Figure S2:**
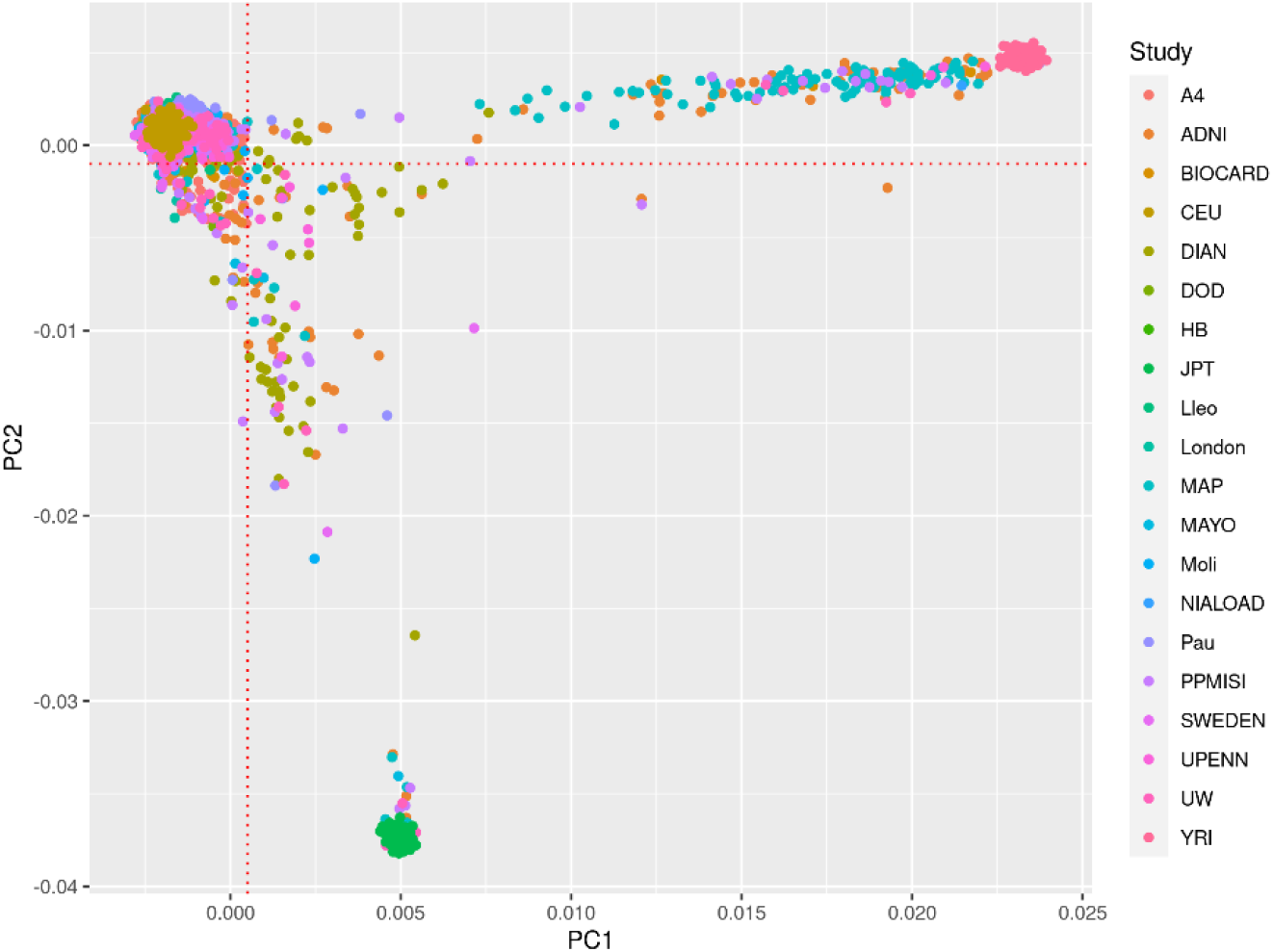
First two principal components of the genetic population structure across 17 cohorts together with the reference HapMap data (CEU, JPT, and YRI). First two principal components of the genetic population structure across 17 cohorts (A4, ADNI, BIOCARD, DIAN, DOD, HB, Lleo, London, MAP, MAYO, Moli, NIALOAD, Pau, PPMISI, SWEDEN, UPENN, and UW) analyzed in this study, together with reference HapMap data (CEU, JPT, and YRI). The red dotted lines represent the thresholds (PC1 < 0.0005 and PC2 > -0.0010) for defining 9526 Northwestern European subjects. Abbreviations: PC, principal component.

**Figure S3:**
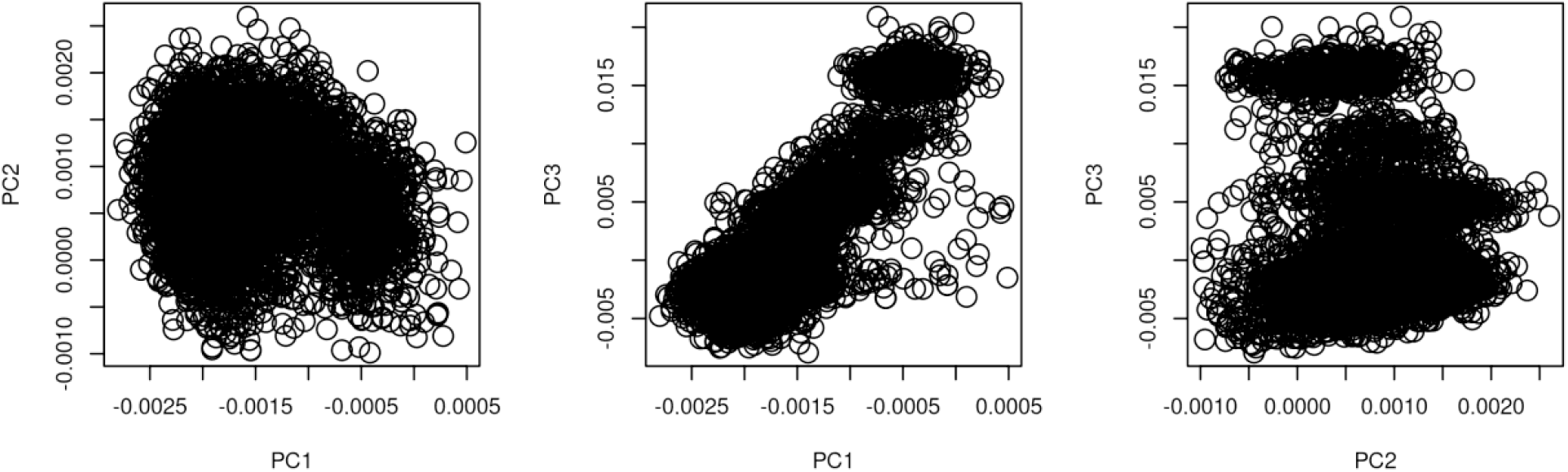
First three principal components of the genetic population structure in Northwestern European subjects across all 17 cohorts. First three principal components (PCs) of the genetic population structure across 17 cohorts (A4, ADNI, BIOCARD, DIAN, DOD, HB, Lleo, London, MAP, MAYO, Moli, NIALOAD, Pau, PPMI, SWEDEN, UPENN, and UW) analyzed in this study. Abbreviations: PC, principal component.

**Table S1.** Demographic characteristics for each analyzed cohort.

| Cohort name | Participants |  |  | Diagnosis % (AD/CN) |  |  | Age, mean (sd) |  |  | Sex % (F/M) |  |  | APOE4+ (%) |  |  |
| --- | --- | --- | --- | --- | --- | --- | --- | --- | --- | --- | --- | --- | --- | --- | --- |
|  | All | 60-80 | CN | All | 60-80 | CN | All | 60-80 | CN | All | 60-80 | CN | All | 60-80 | CN |
| MAP | 1323 | 954 | 565 | 27/61 | 29/59 | 0/100 | 70 (10) | 71 (5) | 70 (5) | 54/46 | 53/47 | 55/45 | 41 | 44 | 35 |
| NIALOAD | 24 | 15 | 14 | 17/79 | 7/93 | 0/100 | 74 (9) | 72 (5) | 71 (5) | 62/38 | 60/40 | 57/43 | 21 | 27 | 29 |
| ADNI | 1734 | 1368 | 401 | 63/27 | 61/29 | 0/100 | 74 (7) | 72 (5) | 71 (5) | 45/55 | 46/54 | 56/44 | 44 | 46 | 29 |
| BIOCARD | 215 | 97 | 53 | 0/69 | 0/55 | 0/100 | 58 (10) | 65 (5) | 64 (4) | 60/40 | 53/47 | 60/40 | 33 | 30 | 26 |
| DIAN | 411 | 17 | - | 8/6 | 0/0 | - | 39 (11) | 63 (3) | - | 56/44 | 53/47 | - | 30 | 41 | - |
| HB | 102 | 71 | - | 100/0 | 100/0 | - | 68 (9) | 70 (6) | - | 45/55 | 2/58 | - | 53 | 56 | - |
| Lleo | 137 | 81 | 39 | 36/64 | 52/48 | 0/100 | 64 (10) | 70 (6) | 68 (4) | 65/35 | 64/36 | 64/36 | 36 | 41 | 31 |
| London | 249 | 192 | 18 | 27/8 | 26/9 | 0/100 | 69 (9) | 70 (6) | 65 (4) | 46/54 | 46/54 | 28/72 | 47 | 44 | 33 |
| Moli | 232 | 155 | 31 | 56/22 | 56/20 | 0/100 | 65 (8) | 68 (6) | 68 (6) | 63/37 | 59/41 | 39/61 | 37 | 37 | 16 |
| Pau | 143 | 106 | - | 12/0 | 12/0 | - | 67 (9) | 70 (5) | - | 50/50 | 47/53 | - | 36 | 35 | - |
| MAYO | 400 | 219 | 181 | 6/76 | 4/83 | 0/100 | 79 (6) | 76 (3) | 76 (3) | 38/62 | 42/58 | 43/57 | 28 | 31 | 27 |
| SWEDEN | 284 | 194 | - | 0/0 | 0/0 | - | 75 (8) | 74 (5) | - | 63/37 | 58/42 | - | 76 | 77 | - |
| UPENN | 113 | 80 | - | 0/0 | 0/0 | - | 72 (9) | 72 (5) | - | 58/42 | 51/49 | - | 58 | 68 | - |
| UW | 355 | 197 | - | 0/0 | 0/0 | - | 63 (16) | 70 (6) | - | 50/50 | 49/51 | - | 43 | 48 | - |
| PPMISI | 546 | 316 | 87 | 0/29 | 0/28 | 0/100 | 62 (10) | 68 (5) | 68 (5) | 33/67 | 29/71 | 28/72 | 27 | 25 | 26 |
| A4 | 3106 | 2935 | 2934 | 0/100 | 0/100 | 0/100 | 71 (5) | 71 (4) | 71 (4) | 60/40 | 61/39 | 61/39 | 36 | 37 | 37 |
| ADNIDOD | 152 | 147 | 98 | 32/68 | 33/67 | 0/100 | 69 (5) | 69 (4) | 69 (4) | 1/99 | 1/99 | 1/99 | 27 | 27 | 30 |

Demographics of subjects at the time of amyloid PET imaging and CSF sampling across each cohort. This table summarizes the demographic characteristics of participants for each analyzed cohort. Participant stratification was performed at three levels, 1) All, All of the subjects; 2) 60-80, Subjects that are 60 to 80 years old; 3) CN, Subjects that are controls and aged 60 to 80 years. For each cohort, we report number of subjects in each stratum, their diagnosis (number of AD and controls subjects), mean age of the participants and standard deviation in the age across cohort, percentage of females and males, and percentage of APOE4+ subjects. Abbreviations: %, percentage; AD, Alzheimer's Disease; CN, Controls; sd, standard deviation; F, Females; M, Males; APOE4+, *Apolipoprotein E4* positive; All, All participating subjects (controls and AD); 60-80: Participating subjects aged 60-80 (controls and AD); CN: 60-80, Control participating subjects aged 60-80; MAP, Memory and Aging Project (MAP); NIALOAD, National Institute on Aging Late Onset Alzheimer's Disease; ADNI, Alzheimer's Disease Neuroimaging Initiative; BIOCARD; DIAN, the Dominantly Inherited Alzheimer Network; HB; Lleo; London; MOLI; Pau; MAYO, Mayo Clinic; SWEDEN; UPENN; UW; PPMI, Progression Markers Initiative; A4, Anti-Amyloid Treatment in Asymptomatic Alzheimer's Disease; ADNIDOD, ADNI Department of Defense.

**Table S2.**
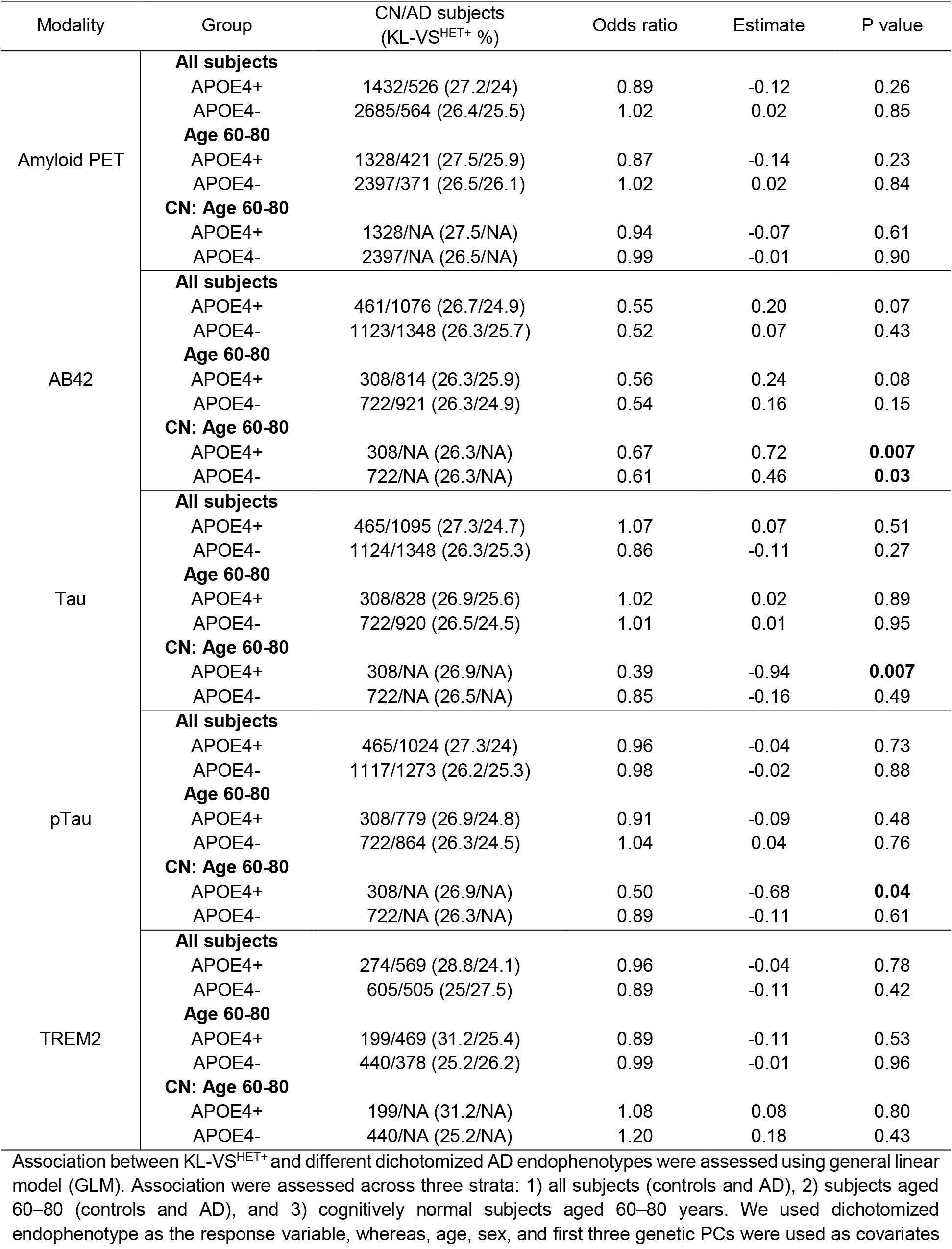
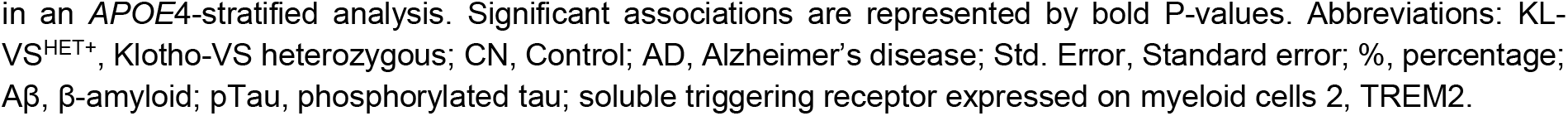
Genetic association of KL-VS^HET+^ with dichotomized AD endophenotypes (GLM), stratified by APOE4 status.

**Table S3.** Genetic association of KL-VS^HET+^ with AD endophenotypes in cognitively normal age- and sex-matched subjects, stratified by APOE4 status.

| Modality | Group | CN subjects<br>(KL-VS<sup>HET+</sup> %) | Odds ratio | Estimate | P value |
| --- | --- | --- | --- | --- | --- |
| Amyloid PET | APOE4+ | 1324 (27.6) | 0.93 | -0.07 | 0.57 |
|  | APOE4- | 1324 (25.5) | 0.95 | -0.05 | 0.72 |
| Aβ42 | APOE4+ | 307 (26.1) | 0.68 | 0.75 | **0.005** |
|  | APOE4- | 307 (26.1) | 0.67 | 0.70 | **0.03** |
| Tau | APOE4+ | 307 (26.7) | 0.39 | -0.93 | **0.008** |
|  | APOE4- | 307 (26.1) | 0.49 | -0.71 | 0.11 |
| pTau | APOE4+ | 307 (26.7) | 0.51 | -0.67 | **0.04** |
|  | APOE4- | 307 (26.1) | 0.75 | -0.28 | 0.45 |
| TREM2 | APOE4+ | 192 (29.7) | 1.15 | 0.14 | 0.66 |
|  | APOE4- | 192 (23.4) | 1.08 | 0.08 | 0.82 |

Association between KL-VS<sup>HET+</sup> and different dichotomized AD endophenotypes were assessed using general linear model (GLM). We used dichotomized endophenotype as the response variable, whereas, age, sex, and first three genetic PCs were used as covariates in an *APOE4*-stratified analysis. For each modality, we considered equal number of age- and sex-matched participants from both *APOE4* strata. Significant associations are represented by bold P-values. Abbreviations: KL-VS<sup>HET+</sup>, Klotho-VS heterozygous; CN, Control; AD, Alzheimer's disease; Std. Error, Standard error; %, percentage; Aβ , β-amyloid; pTau, phosphorylated tau; soluble triggering receptor expressed on myeloid cells 2, TREM2.

